# The Unreliability of *Estimated Release Dates* in Hospital Drug Shortage Management: A Case Study of Hospital Pharmacy Operations During the COVID-19 Pandemic

**DOI:** 10.1101/2025.07.10.25331166

**Authors:** Noah Chicoine, Jacqueline Griffin

## Abstract

Drug shortages are prominent, persistent operational challenges that hospital pharmacies have been facing for years. During a drug shortage, hospital pharmacists must solve the problem of how best to invest resources to mitigate the effect of the drug shortage on patient health care. One piece of data they use to inform their decision-making is the estimate release date (ERD) of a drug, a point estimate given from the pharmaceutical manufacturer specifying when the next release of a drug (that is on shortage) will occur. Working with a hospital collaborator, we collected a novel set ERD and shipment data to analyze the the accuracy of this information and the impact on decision-making at hospitals. We show via statistical analysis that ERD information tends to be an inaccurate indicator of when the hospital should expect to receive more product and is subject to change randomly and at random intervals, adding additional complexity to managing drug shortages. We discuss managerial insights that stem from this analysis and lay a foundation for future research studying decision-making with unreliable ERD information.

## 1. Introduction

Drug shortages are prominent and severe events that have regularly affected hospital pharmacy operations and patient health care for decades [1]. The adverse effects of these shortages on patient health care outcomes have been documented extensively in the literature [2; 3]. Drug shortages at hospitals have delayed patient surgeries, chemotherapy treatments, and caused adverse side effects when alternative products have been utilized to treat patients with critical needs. Additionally, drug shortages have increased the likelihood of medicine administration errors from medical staff when administering alternative products [4]. To prevent these events and maintain the quality of care, some hospitals have established drug shortage management protocols and implemented software solutions to help manage communication and inventory during a shortage. These protocols and tools, however, are only part of the laborious process of managing drug shortages, which involves extensive internal coordination, problem-solving, and communication, all aimed at preventing shortages from directly affecting patient care.

During the COVID-19 pandemic, many drug products used to treat COVID-19, like neuromuscular blockers, antiviral medications, and corticosteroids, were in short supply at hospitals all over the United States [5; 6]. Additionally, COVID-19 disrupted pharmaceutical manufacturing operations at several manufacturing sites, affecting other drug products not directly related to COVID-19 treatment. COVID-19 created a perfect storm for drug product management in hospitals because increased patient demand was coupled with a limited drug supply, caused by fragility in pharmaceutical supply chains [7]. This presented a unique opportunity to collect vast amounts of temporary data related to drug shortage management, including shortage management strategies, pharmacists’ concerns and stress, and changes in medical operations at hospitals.

To take advantage of this opportunity, our research group conducted a case study with research collaborators from from a large U.S. hospital system throughout the COVID-19 pandemic. Through this collaboration, we discussed how pharmacy managers monitor and prioritize mitigating drug shortages on a daily basis, common mitigation strategies, information collected from stakeholders, and stories about ongoing shortages. Additionally, we discussed the information the pharmacy supply chain managers received from upstream suppliers (i.e., wholesalers and manufacturers) to gain a better understanding of the information environment in which they worked and to potentially gain insight into the operations of manufacturers and wholesalers during drug shortages. One prominent theme that emerged as a research area of interest was the operational challenges related to the resupply information they received from upstream stakeholders in the supply chain. Below, we summarize the major takeaways from discussions with our hospital collaborators and introduce the operational problems presented by resupply information.

### 1.1 Hospital Operations Challenges During Drug Shortages

Our collaborators described the following workflow as a typical strategy when managing drug shortages. During a shortage, our collaborators evaluate their product inventory using both inventory management software and manual assessments of inventory throughout the hospital. Additionally, they assess the status of future supply. They determine when they expect to receive more product and whether substitute products are available. Finally, with this knowledge, pharmacy management decides which operational solutions to implement to mitigate the effects of the shortage on patient care, considering the solution’s cost and the likely disruption to hospital operations. Examples of mitigation strategies are shown below in Table 1, along with their purpose and related costs. These strategies align with previous literature studying drug shortage management at other US hospitals [8].

**Table 1:** Drug shortage mitigation strategies utilized at a major US hospital system during the COVID-19 pandemic.

| Mitigation Strategy | Purpose | Related Costs |
| --- | --- | --- |
| Compounding drugs in-house | Synthesize more vials/bags of a drug from smaller volume presentations | Pharmacy staff labor |
| Extra consultation with physicians | Determine possible alternative medications and communicate the shortage status | Pharmacy and physician time and labor |
| Updating prescription software to reflect low inventory | Persuade physicians to use alternative medications when possible | Pharmacy and physician time and labor |
| Contact the manufacturer or wholesaler directly | Inquire about the drug product shortage status and/or request emergency or alternative shipments of available product | Pharmacy time and labor |
| Switch practices to alternative medication | Treat patients when little or no inventory or the preferred product is left | Nursing staff training, pharmacy time and labor, sub-optimal patient care, increased medication costs |

As previously mentioned, the pharmacy management must make decisions to implement one or more mitigation strategies based on their current supply, the status of future supply, and the impact on the hospital and its patients. In some cases, manufacturers may communicate *estimated release dates* (ERDs) to supply chain partners, which hospital pharmacies may use to inform their decision-making. As implied by the name, ERDs are dates by which pharmaceutical manufacturers expect to release the next batch of a drug into the supply chain. During a drug shortage, hospitals often do not receive any product from the wholesaler or manufacturer for extended periods of time because there is limited stock left in the supply chain. Thus, an ERD is often synonymous with the next time stock will be available to fulfill a hospital’s pending order. Hospital pharmacists can use ERD information to assess whether they can sustain a healthy supply throughout a shortage or whether they should consider implementing mitigation tactics. Although they estimate the time of the next shipment, ERDs do not communicate how much product will be released or what quantity hospitals can expect to receive.

While pharmaceutical manufacturers are not required to disclose this information, some voluntarily communicate ERDs. When they have done so in the past, wholesalers collected, aggregated, and reported the information to hospitals in a weekly report that listed all the drugs currently in short supply and the ERDs if they were provided. This flow of information through the supply chain is shown in Figure **??**. Recently, wholesalers have switched from sending ERDs in aggregated reports to posting them on their online ordering sites individually for each drug product.

A key challenge of utilizing ERD information to manage drug shortages is that ERD information is often unreliable. Our collaborators have described on multiple occasions how ERD information from manufacturers has not accurately indicated when shipments would arrive at the hospital. Additionally, ERDs are uncertain and subject to change, even at the last minute.

When ERD information is inaccurate, it can lead hospital pharmacists to make inappropriate inventory management decisions, which can result in the hospital incurring additional, avoidable operational costs, in the form of time, money, and labor (see Table 1). More specifically, inaccurate ERD information can cause two unique problems that result in increased costs of managing drug shortages. First, if an ERD is far away, hospital pharmacies may implement proactive mitigation measures. However, if the shipment arrives earlier than expected, those mitigation measures will be rendered unnecessary, need to be reverted, and will have cost the hospital extra time, money, and labor.

On the other hand, if an ERD is soon, the pharmacy team may determine that the hospital can maintain its current healthcare practices until the shipment arrives. However, if the shipment arrives later than expected, the hospital and pharmacy team may have to implement widespread mitigation measures very quickly, which is costly for the hospital and stressful for hospital staff, more so than if these measures had been implemented earlier.

In summary, ERD information adds complexity to drug shortage management problems when the ERD does not accurately communicate a shipment date. Not only do pharmacists need to discern the best mitigation strategies to combat drug shortages and preserve patient healthcare quality, but they also have to choose how much trust to place in the ERD information provided to them and how to act accordingly. ERD information presents a unique inventory management problem that involves information sharing, patient costs, and monetary costs, which is not discussed in the drug shortage literature but is becoming a standard part of hospital pharmacy practices.

### 2.1 Problem Statement

Managing drug shortages is a challenging task for hospital pharmacies to navigate in a way that preserves patient health outcomes and ensures operational efficiency at the hospital. Many of the circumstances surrounding drug product shortages, such as patient demand, ERD information, and shipping times, are out of the hospital’s control. The primary thing hospital pharmacies have control over is their internal response to the shortage, including mitigation strategies they can implement. The inaccuracy and uncertainty of ERD information make shortage management particularly challenging because they directly affect the timing and resulting costs of shortage mitigation strategies. A hospital may make a cost-effective mitigation decision, but an ERD change can completely undermine the viability of the original plan and cause hospitals to incur additional operational costs. Thus, a primary focus of this study and future research is to find ways to improve shortage mitigation operations at hospitals by improving inventory and ordering decisions based on current information-sharing practices.

This study has two distinct aims. The first aim is to statistically assess the accuracy and variability of ERD information for both managerial insights and scientific novelty. To our knowledge, the data set presented in this study is the first instance of pharmaceutical ERD information data in the literature. The analysis of this data will provide a basis for future research of lead time information sharing in pharmaceutical supply chains (and other contexts) and also provide hospitals with insights for future drug shortage management decision-making. Thus, for the first aim of this study, we address the following research questions:

**RQ1.1**. Do ERDs accurately portray when shipments will arrive at a hospital? If an ERD is updated, does it tend to be more accurate than the previously reported ERD?

**RQ1.2**. Does ERD accuracy differ between manufacturers, drug classes, or product packaging types?

The second aim of the study is to provide an understanding of how ERDs and their accuracy evolve, particularly in the lead-up to shipment arrival. Quantifying how ERDs change over time in this study will help lay the foundation for developing inventory management models in future studies. With an accurate model of how ERDs evolve, future studies can explore optimal inventory management strategies in the context of drug shortages. To achieve this second aim, the following research questions are addressed:

**RQ1.3**. How frequently are ERDs updated? Are the changes memoryless (i.e., inter-arrival times are geometrically distributed)?

**RQ1.4**. By how much do ERDs change? Does the magnitude of the change depend on the current ERD?

In the following sections, we discuss the current state of the literature surrounding lead time information in pharmaceutical supply chains, the methods we use to analyze the data and quantify trends, and present the findings and conclusions from this work.

## 2. Literature Review

To our knowledge, this is the first study to collect and analyze real-world lead time data from pharmaceutical supply chains. Though there is an abundance of work studying other elements of drug shortages, such as causes, effects, management strategies, and prevention strategies [1], there is a lack of (or shortage of) scientific literature that analyzes ERD information or any form of lead time information data in pharmaceutical supply chains [9]. Burinskiene (2019) and Chen et al. (2021) discuss the concepts of lead times in a pharmaceutical context, but do not provide any data or analysis of lead time information in practice [10; 11]. This gap in the literature is present despite the National Academies Committee on the Security of America’s Medical Product Supply Chain highlighting information sharing and transparency as key components to pharmaceutical supply chain fragility [12]. One study by Yongsatianchot et al. examines human decision-making in an inventory management scenario with ERD information [13]. This study, however, does not include any empirical data or analysis of real ERD information from a pharmaceutical supply chain, nor does it discuss the operational costs associated with inaccurate ERDs. Our study builds on the work by Yongsatianchot and colleagues by providing evidence of the kinds of uncertainty real-world decision-makers experience in practice. Additionally, we aim to set the foundations for future work in operations management that examines the problem of uncertain lead times in drug shortage contexts, a unique context in the broader supply chain research space.

Despite limited research on lead time information sharing in pharmaceutical supply chains, other studies have examined the impacts of this information in generic supply chain contexts. Research has shown that lead time information can be valuable to a firm implementing base stock inventory policies, particularly in scenarios where lead times are highly variable [14; 15] or high shipments are high volume [16]. Dobson and Pinker (2006) used a queuing model to show that a firm sharing lead time information with its customers can be beneficial to stakeholders throughout the supply chain [17]. Altug et al. (2011) developed a state-dependent base stock inventory policy that accounts for advanced capacity information from upstream suppliers and demonstrated how this advanced information can improve supply chain performance [18]. Additionally, Jain et al. (2017) examined how knowledge of supplier inventory impacts optimal retailer inventory policies in a multi-agent supply chain game [19].

Pharmaceutical supply chains pose unique risks and complexities compared to other supply chains, as they frequently experience disruptions [20] and serve critical patient needs [8]. Thus, a shift in context (one that includes disruptions and high backlog costs) is necessary to model these systems appropriately. There is a large body of literature that examines inventory management during supply chain disruptions [21]. Czerniak et al. (2024) developed adaptive inventory models as a means of addressing recurrent and chronic drug shortages [22]. Rosovski et al. (2020) discuss mitigation strategies hospitals employed proactively against Heparin shortages that ensued from swine flu in 2019 [23]. However, these studies do not address lead time variability or uncertainty, which are standard parts of pharmacy inventory management.

To our knowledge, there are only a few studies that combine the concepts of lead time information with the context of supply disruptions. Mehrotra and Schmidt (2021) use stochastic linear programming and data from a manufacturing firm to show the value of different types of lead time information, including ranges, distributions, and accurate points [24]. Jin et al. (2024) develop a dynamic programming framework to determine how best to mitigate operational costs when given upstream information about the anticipated disruption duration [25]. Both of these studies provide a strong foundation for developing inventory management models in the context of drug shortages. However, before that is done, it is essential to capture a nuanced representation of ERD accuracy and uncertainty for such a model. Our study conducts a deep data analysis of ERD information gathered from a hospital collaborator to provide a basis of understanding of ERD information accuracy and trends. The results of this study can serve as a foundation for future models adapted from previous studies, like those mentioned above.

## 3. Methods

### 3.1 Data Collection & Cleaning

The ERD and shipment data analyzed in this study were compiled from two sets of reports collected from October 2022 to June 2023. The ERD data came from weekly ERD reports sent to our hospital collaborators from their wholesaler. The ERD reports featured hundreds of unique products, only some of which the hospital purchased regularly. Over time, the number of drug products included on the ERD reports increased, as seen in Figure **??**. This trend aligns with reports of increasing drug shortages noted in 2020 by the FDA [26] and ASHP [27].

The shipment data came from daily shipment arrival reports generated by the hospital. These reports contained short lists of drug products that arrived at the hospital the previous day. In total, 156 shipment arrival reports were collected. Some days were missing in the dataset because there were days when the hospital did not generate shipment arrival reports, and the reports could not be generated retroactively. Since each report contained very few shipment entries, most of which were for NDCs not included in the ERD reports, it is assumed that missing shipment arrival reports had a minimal impact on the dataset or the calculated accuracy of ERDs.

To combine the data sets for analysis, we modified the arrival dates in the shipment arrival reports so that all drugs arriving the same week (Sunday to Saturday) were labeled with the same date (Monday), which matched the day the hospitals received the ERD reports. The analysis was then conducted on the scale of weeks. Additionally, extensive data cleaning and filtering procedures were performed to identify the subset of drug products that were included in both the ERD reports and had ERD information, and the shipment arrival reports. From this cleaning procedure, we found *n* = 197 ERD-shipment pairings, referred to as “ERD trajectories” throughout the remainder of this paper. The 197 ERD trajectories represent 122 unique NDCs.

For the analysis, the dataset was parsed into multiple subgroups to compare ERD accuracy between these groups. More specifically, the dataset was broken down into groups based on ERD magnitude, manufacturer, drug class ([28]), and packaging type to examine differences in statistical tendencies within these categories. These categories were chosen because we hypothesized that ERD accuracy could be linked to the magnitude of the original ERD, the manufacturing and communication practices of individual manufacturers, or the sourcing of ingredients and materials. So, ERD trajectories were grouped by ERD magnitude and each unique manufacturer, drug class, and packaging type, except for groups that had 10 or fewer trajectories. These groups were combined into an “other” group in their respective categories to maintain a sufficiently large sample size for statistical analysis. The resulting groupings are outlined below in Table 2. The manufacturer groups, specified by individual unique manufacturers, were labeled as “Man. 1”, “Man. 2”, etc. to maintain anonymity.

**Table 2:** Data subgroups used in analysis.

| Category | Subgroups | $n$ |
| --- | --- | --- |
| **ERD Magnitude** | 1) 1-5 weeks | 96 |
|  | 2) 6-10 weeks | 48 |
|  | 3) 11-15 weeks | 14 |
|  | 4) 16-60 weeks | 14 |
| **Manufacturer** | 1) Man. 1 | 68 |
|  | 2) Man. 2 | 58 |
|  | 3) Man. 3 | 19 |
|  | 4) Man. 4 | 14 |
|  | 5) Man. 5 | 11 |
| | 6) Other ($n_i \leq 10$) | 27 |
| **Drug Class** | 1) Amide local anesthetic | 72 |
|  | 2) Nucleoside metabolic inhibitor | 18 |
| | 4) Other ($n_i \leq 10$) | 107 |
| **Drug Packaging** | 1) Multi-dose vials | 82 |
|  | 2) Single-dose vials | 82 |
|  | 3) Bags | 23 |
|  | 4) Ampules & syringes | 10 |

### 3.2 Measurements & Statistical Methods

To address the research questions posed in Section 1.1.2, we examined ERD accuracy and ERD updates in a variety of ways. First, we measured ERD accuracy by calculating the “shipment lateness” of each ERD trajectory in the dataset, a measure of how much later the shipment arrived after the original ERD specified it would. We examined shipment lateness across the whole dataset and also compared shipment lateness between subgroups specified in Table 2. Since the ERD data is discrete (measured in weeks) and strictly positive, violating key assumptions of parametric tests, we utilized non-parametric techniques to test for statistical significance within and between subgroups of the dataset. Specifically, we used Wilcoxon Signed-Rank (WSR) tests, Mann-Whitney U (MWU) tests, and Kruskal-Wallis (KW) tests [29] for one-sample, two-sample, and multi-sample comparisons, respectively.

Then, we assessed how ERDs change over time by calculating three measures for each ERD trajectory: (1) the number of ERD updates that occur before the shipment arrives, (2) the inter-arrival time of each ERD update in each trajectory, and (3) the magnitude of each ERD update. This assessment provides a foundation for modeling pharmacy inventory management with this uncertain lead time information. We do not perform any statistical testing on the number of ERD changes in each trajectory. However, we did fit the inter-arrival time measures against a geometric distribution to test whether ERD updates follow the memoryless property. Before fitting, inter-arrival times that exceeded the 99.5 percentile of a geometric distribution following the mean of the dataset were removed. Then, a χ^2^ test was used to test the fit of inter-arrival times of ERD updates against a geometric distribution. Additionally, a KW test was used to assess if updated ERDs were more accurate than previously communicated ERDs. Finally, we examined the distribution of ERD update magnitudes that occurred across all ERD updates in the dataset. To assess if the magnitude of the updates was dependent on the original ERD, we used a KW test across distributions of ERD updates where the original ERD was between 1 and 20 weeks.

## 4. Results

Figure **??**a shows a box plot of shipment lateness for all 197 ERD trajectories. A shipment lateness close to 0 in these plots is akin to an ERD being “accurate”. The results of a WSR test found that shipments typically arrived significantly later than the communicated ERD (*p* < 0.001, *H*_*a*_ : *m* > 0). The box plot also depicts the wide variance in shipment arrivals relative to the ERD, which augments testimonies from hospital pharmacy staff that ERDs do not reliably approximate when a shipment will arrive.

The box plots of shipment lateness broken down by original ERD magnitude are shown in Figure **??**b. A KW test found that these distributions differed significantly, suggesting that ERD accuracy may depend on how far away the ERD is from the time it’s reported. Upon further investigation, MNU tests showed that ERDs in the 11-15 week range differ in accuracy compared to ERDs in the other groups (*p* < 0.05). WSR tests found ERDs in the 11-15 week range were not significantly ahead of the shipment time (*p* = 0.966). However, ERDs were significantly ahead in the other groups, which is consistent with the results for the entire dataset (Figure **??**a).

We conducted a similar analysis on the subgroups of the data specified in Table 2. Figure **??** shows box plots of shipment lateness broken down by the groups shown in Table 2. For most groups, WSR tests found that ERDs were significantly ahead of the shipment time, except for Manufacturer 4, Manufacturer 5, nucleoside metabolic inhibitors, and bag products. These results were supported by MWU tests, showing that the groups differed significantly within their categories. The results of these statistical tests and the tests discussed above are summarized below in Table 3.

**Table 3:**
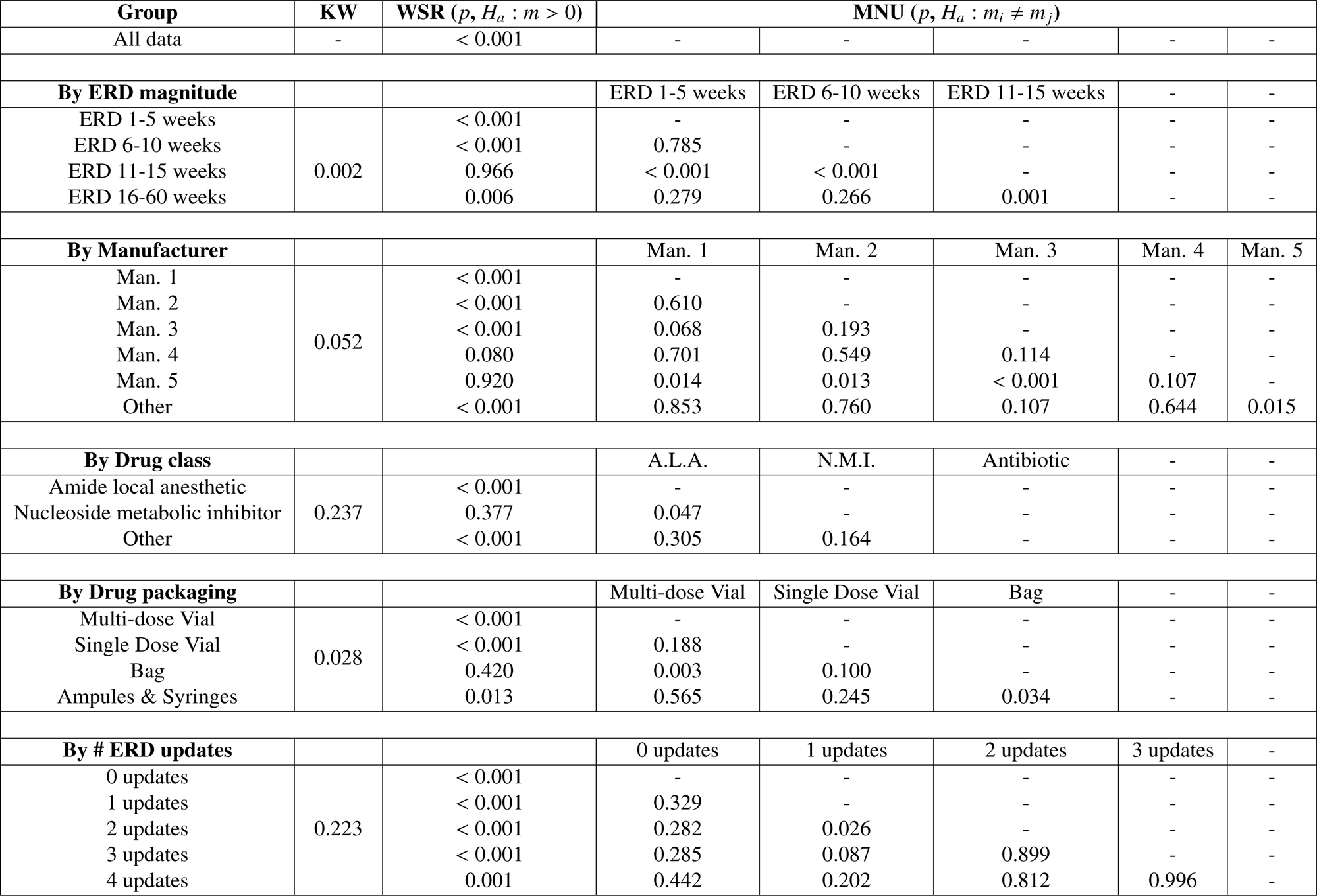
Statistical test results by subgroups.

Second, we measured how often ERDs change within ERD trajectories, and whether the shipment lateness of updated ERDs improves significantly compared to past ERDs. Figure **??**a shows a histogram of the number of ERD updates in each ERD trajectory in the dataset. The figure highlights how almost half of the trajectories (48.7%) featured two or more ERD updates before the shipment arrived, with some featuring as many as 10 to 12 updates. Figure **??**b shows the distribution of inter-arrival times of these ERD updates. After removing the outliers, a χ^2^ test found that the distribution of inter-arrival times was not significantly different than a geometric distribution (*p* = 0.11), indicating that the time between ERD updates could be modeled as a random Poisson process. Correspondingly, as a Poisson process is inherently memoryless, the likelihood of future ERD updates in this dataset is independent of past updates.

Figure **??**c shows box plots of shipment lateness initially, and after the first, second, third, and fourth updates. Similarly to the entire dataset, each group was still significantly ahead of the shipment arrival (*p* < 0.05). Additionally, a KW test concluded that these distributions do not significantly differ from one another (*p* = 0.223), suggesting that ERD accuracy does not statistically improve as ERDs are updated. These findings are supported by pairwise comparisons of consecutive updates, shown in Table 3.

Lastly, we measured the magnitude of the ERD updates in the dataset and determined whether the updates depend on the current ERD. In the dataset, ERDs changed 18.9% of the time week to week. Figure **??**a shows a histogram of the magnitudes of these updates when they occurred. ERDs were pushed back to a later date 71% of the time and changed to a closer date 29% of the time, with an overall average change of 2.58 weeks. Figure **??**b shows box plots of the magnitude of the ERD updates grouped by how far away the current ERD is. A KW test suggests that these distributions of changes do not differ significantly (*p* = 0.101), further indicating that the magnitude of ERD updates does not statistically depend on the current ERD itself. Thus, the magnitude of change of an ERD is independent of the current ERD.

## 5. Discussion

The focus of this study was to examine ERD and hospital shipment data statistically to (1) provide managerial insights that could improve drug shortage management decision-making at hospitals, and (2) quantify ERD accuracy to provide a foundation for modeling drug shortage management with uncertain lead time information.

With respect to managerial insights, the results above indicate a persistent issue in drug shortage management. Pharmacists face an ongoing challenge in making shortage mitigation decisions for multiple shortages every day [8]. These decisions have direct consequences on clinical health care and hospital operations and are reliant on ERD information. As the results show, most of the time, this information is not trustworthy for operational decision-making. The data and statistical tests used in this study revealed that communicated ERDs are typically earlier than the actual shipment date, with a wide variability in overall accuracy. Even for a subset of products, like those from Manufacturer 5 and for bag products, which are the most reliable, it is not uncommon for individual shipments to arrive several weeks earlier or later than the specified ERD. Additionally, when ERDs are updated, the updates occur at random intervals and with a wide range of magnitude, making it difficult to anticipate when future updates will occur and what the next ERD will be. An updated ERD is also not necessarily more accurate than the previously communicated ERD. This statistically random evolution of ERDs could be attributed to many different external factors, including variable shipment lead times upon release, wholesalers not fulfilling every hospital’s back orders upon each release, or the difficulty involved in manufacturers accurately predicting the date of future releases. Further investigation is needed to discern the causes. Regardless, with the need for pharmacists to be risk-averse when acquiring life-saving medication during drug shortages, there is reason to be skeptical about when the next shipment of a product will truly arrive.

The current state of inaccurate lead time information during drug shortages creates a difficult problem for pharmacy managers. ERD information is directly relevant in managing drug shortages in hospitals as it is a primary indicator of when the hospital should expect to receive more product. It seems counterintuitive that pharmacy managers should ignore this information, but currently, they must rely on their perceptions of how trustworthy ERD information is when making decisions.

Given the results of this study, it may be best in practice to seek out alternative information on future shipment arrival time or weight ERD information low among factors that inform mitigation strategy implementation at the hospital. Hospital pharmacy managers may contact sales representatives directly to get more information to contextualize the ERD. There may be value in gathering additional information about supply availability from other third-party sources. However, in a setting already limited in resources, this strategy creates an additional challenge of choosing how and where to invest those limited resources in obtaining better-quality information. This adds another managerial problem to the overall challenge of managing drug shortages. As an alternative to investing in more information, it may be more effective to prioritize other factors such as the costs of mitigation strategies and the potential for avoiding patient harm. This could be done on a case-by-case basis, where ERDs are weighted higher for specific subsets, like nucleoside metabolic inhibitors and bag products, for which ERDs tend to be more accurate.

### 5.1 Limitations

One limitation of this study is that the dataset was collected from only one hospital. ERD evolution and shipment lateness might present differently at other hospitals due to differences in supply chain structures. Likely, hospitals would all receive similar ERD information from manufacturers. However, differences in wholesaler contracts, geography, and ordering behavior could impact when shipments arrive at a hospital. Thus, the reliability and accuracy of ERD information may differ between hospitals. Additionally, due to hospitals having unique formularies, patient profiles, specializations, uses, and inventory management strategies, the same ERD information may affect their decision-making differently. Thus, examining ERD at a broader scale across hospitals has the potential to yield more system-wide insights and could reveal additional needs or inventory management problems that arise from ERD information. The wide variability in shipment lateness across different manufacturers may suggest that inaccurate ERDs could be affecting other hospitals, but additional data collection is needed to confirm this.

Additionally, this dataset was collected at a time when pharmaceutical supply chains were recovering from years of COVID-19-related disruptions. External economic factors, such as global supply chain disruptions, may have contributed to manufacturers’ and wholesalers’ lead time estimations. Additional data is needed to examine whether this trend in ERD information has changed since the COVID-19 pandemic.

### 5.2. Future Directions

There are opportunities for quantitative modeling based on the insights gained from this study. First, human decision-making and information systems models can be developed to explore the tradeoffs of acquiring more accurate resupply information given the financial and operational constraints of hospital systems. These models are needed to investigate what other information may be worth acquiring to aid decision-making in the high-stakes scenarios of drug shortage. These types of models could be used to aid ongoing decision-making in hospital settings, given the current information-sharing infrastructure.

Second, operations management models can be developed to investigate optimal drug shortage management strategies given the characteristics of ERD information discovered in this study. The results of this study show that ERD updates occur at independent, geometrically random intervals and that the newly updated ERD does not statistically depend on the previously communicated ERD. Additionally, the accuracy of an ERD is not statistically dependent on these updates and is likely to remain early after each update. These are useful characteristics that can be incorporated as assumptions in stochastic models to gain insights into optimal inventory management strategies in the pharmaceutical supply chain environment. Models like these could be used to develop assistive tools for managing drug shortages and test changes to the information-sharing infrastructures within pharmaceutical supply chains for efficacy. For example, these models could be used to assess the cost savings of implementing *ERD ranges* in place of ERD point estimates. Current supply chain literature suggests that ERD ranges, or potentially other forms of upstream inventory transparency, could be beneficial for hospital operations costs while managing drug shortages [19; 24]. However, models that are more nuanced towards the unique traits of pharmaceutical supply chains, such as high cost of backlog and high costs of alternative suppliers and products, are needed.

## 6. Conclusions

Drug shortages are common and chronic problems that hospitals constantly have to navigate [30; 8; 20]. Hospital pharmacies use all available resources and information to make informed decisions about inventory management and medical practice, ensuring patient safety during shortages. ERD information, although it has not yet been discussed or studied in the scientific literature, is a crucial piece of data regarding the upstream stability of future drug supplies. We show, using a novel dataset from a large US hospital, that ERD information tends to be an uncertain and unreliable estimate of the future shipment arrivals. In addition to exploring this data for managerial insights, we also laid the foundation for future models of pharmacy inventory management to incorporate this uncertain lead time information and investigate optimal inventory management policies that take it into account. Further research in the area of information coordination in pharmaceutical supply chains and inventory management in these contexts has the potential to positively impact drug shortage management at hospitals and improve patient care.

## Acknowledgments

This work was funded partially by NSF RAPID Grant #2028449 and NSF SAI Grant #2228510.

## Data Availability Statement

The data used in this study will not be made publicly available due to a nondisclosure agreement between the authors and the collaborating hospital.

